# Spatial Analysis and Multilevel Determinants of Hypertension in Zambia: Analysis of the 2017 WHO STEPS Survey

**DOI:** 10.64898/2026.06.18.26355935

**Authors:** Samuel Mutasha, Simkoko Davies, Chand Nkandu

## Abstract

**Background:** Hypertension is the leading modifiable cardiovascular risk factor globally, with the fastest-growing burden in low- and middle-income countries. This study aimed to estimate national hypertension prevalence, map provincial patterns, assess spatial clustering, and identify individual and community-level determinants among Zambian adults using the 2017 WHO STEPS survey.

**Methods:** This cross-sectional study used data from the 2017 WHO STEPS survey, a nationally representative sample of 4,301 adults aged 18–69 years. Hypertension was defined as systolic BP ≥140 mmHg, diastolic BP ≥90 mmHg, or current antihypertensive use. Spatial autocorrelation was assessed via Moran’s I and LISA. Four nested generalised linear mixed models with PSU-level random intercepts identified individual and community-level determinants.

**Results:** Overall weighted hypertension prevalence was 24.0%. Lusaka recorded the highest prevalence (30.2%), followed by Southern (29.9%) and Muchinga (28.3%) provinces; Western Province had the lowest (12.4%). Spatial clustering was statistically significant but modest (Moran’s I = 0.0247, p < 0.001). Between-cluster variation reduced from ICC = 5.9% to 1.8% in the full model, indicating geographic differences were largely explained by individual characteristics. Age was the strongest predictor; adults aged 60–69 had nearly sevenfold higher odds than those aged 18–29 (AOR 6.92, 95% CI: 4.95–9.66). Women had lower odds than men (AOR 0.64, 95% CI: 0.52–0.79). Obesity (AOR 2.34), overweight (AOR 1.65), high cholesterol (AOR 1.40), diabetes (AOR 1.35), and single marital status (AOR 1.34) were independently significant. Western Province showed consistently lower odds than Central Province (AOR 0.48).

**Conclusion:** Hypertension affects one in four Zambian adults, driven primarily by age, sex, obesity, dyslipidaemia, and diabetes. Geographically prioritised interventions, including community health worker-led screening programmes in Lusaka and Southern Province, would maximise population-level impact. Population-level salt reduction and alcohol policies represent cost-effective complementary strategies. Longitudinal studies with finer spatial resolution are needed to clarify causal pathways underlying observed geographic clustering and inform SDG Target 3.4 progress.

## Introduction

Hypertension, defined as a sustained systolic blood pressure of at least 140 mmHg or a diastolic blood pressure of at least 90 mmHg, is the leading modifiable risk factor for cardiovascular disease and early death worldwide. Over the past three decades, the global burden has doubled, with more than 1.3 billion adults now living with the condition, and the fastest increases seen in low and middle income countries^1^. Uncontrolled hypertension is linked to more than half of all stroke cases and a large share of ischaemic heart disease worldwide, contributing to about 10.8 million deaths each year^1,2^. Although it can be detected and treated at the primary care level, hypertension remains widely undiagnosed and poorly controlled, especially in settings with limited resources^3^.

Sub Saharan Africa now carries a large and rapidly growing share of this burden. The WHO African Region reports the highest age standardised prevalence of hypertension among low and middle income regions ^4^. Africa has seen a two fold increase of adult patients with hypertension to about 390 million due to rapid urban growth, dietary shift to processed and high salt foods, physical inactivity and ageing population ^5,6^. At the same time, detection, treatment, and control remain very low, with only 7.3% of people having their blood pressure under control, leaving many at risk of preventable complications^7,8^.

In Zambia, hypertension is an increasing but still poorly described public health problem. A study using data from the 2017 WHO STEPS survey, reported a prevalence of 18.9% among adults aged 18 to 69 years^9^. However, earlier studies have mostly been based in health facilities or focused on specific areas, which have a limited national representation^10,11^. In addition, the health system at primary care level faces major challenges in managing hypertension, including limited diagnostic tools, shortages of essential medicines, and a lack of trained staff^12,13^.

Addressing hypertension is key to Zambia’s progress toward Sustainable Development Goal target 3.4, which aims to reduce early deaths from non communicable diseases by one third by 2030 ^14^. Progress toward this goal in low income settings will remain slow unless the main drivers of hypertension are clearly identified and addressed^15^. Non communicable diseases now account for nearly a quarter of all deaths in Zambia, with cardiovascular conditions leading, and this burden is expected to increase with ongoing urbanisation and lifestyle change^13,16^.

Despite this growing need, no study in Zambia has combined spatial analysis with a multilevel approach to examine both individual and community level drivers of hypertension. Standard regression methods do not account for the clustering of individuals within communities or the broader context that shapes risk beyond personal characteristics^17,18^. This study therefore uses the 2017 WHO STEPS survey to estimate national prevalence, map provincial patterns, assess spatial clustering, and identify both individual and community level determinants using multilevel logistic regression, providing evidence to support more targeted and equitable interventions in Zambia.

## Materials and Methods

### Study Design, Data Source, and Outcome

This study used individual-level data from the Zambia World Health Organization STEPwise Approach to Surveillance (STEPS) survey, a nationally representative cross-sectional study of adults aged 18–69 years. STEPS collects standardised behavioural (Step 1), anthropometric (Step 2), and biochemical (Step 3) data through a stratified multistage cluster design, with primary sampling units (PSUs) drawn from urban and rural strata across all ten provinces. Data were supplied in Stata format and analysed in R version 4.3. The binary outcome, *Y*_ij_ = 1, indicated hypertension for participant *j* in PSU *i*, defined as a measured systolic blood pressure ≥140 mmHg and/or diastolic blood pressure ≥90 mmHg, or current antihypertensive medication use, consistent with WHO criteria (STEPS variable *highbp2*). All regression models used a logistic link; results are reported as adjusted odds ratios (AORs) with 95% confidence intervals (CIs).

### Analytical Sample and Missing Data

Listwise deletion was applied across all variables included in the full multilevel specification, yielding an analytical sample of *n* = 3,474 complete cases from the total *N* = 4,301 survey participants. Sensitivity of findings to this missingness assumption is noted as a limitation.

### Descriptive Statistics

Participant characteristics were tabulated by hypertension status (Table 1). Survey weights were rescaled to sum to the published STEPS sample size. Categorical associations with hypertension were assessed using design-adjusted Rao–Scott chi-square tests; where more than 20% of expected cells fell below five, Fisher’s exact test with 10,000 Monte Carlo draws was substituted. Waist circumference was summarised with weighted medians and interquartile ranges and compared using a survey-weighted Wald test via *svyglm*. Within-category hypertension prevalence and 95% CIs were estimated using survey-based standard errors.

**Table 1.** Baseline characteristics of study participants stratified by hypertension status, Zambia.

| Characteristic | Level | N<sup>a</sup> | Non-hypertensive n (%) | Hypertensive n (%) | Prevalence % (95% CI) | p-value |
| --- | --- | --- | --- | --- | --- | --- |
| **Total sample (weighted N = 4,301)** |  | 4,301 | 3,269.1 (76.0%) | 1,031.9 (24.0%) |  |  |
| **Age group (years)** |  |  |  |  |  | <0.001 |
|  | 18–29 | 1,520 | 1,319.7 (40.4%) | 200.5 (19.4%) | 13.2 (11.4–15.0) |  |
|  | 30–44 | 1,498 | 1,196.6 (36.6%) | 301.2 (29.2%) | 20.1 (17.9–22.3) |  |
|  | 45–59 | 867 | 571.2 (17.5%) | 295.7 (28.7%) | 34.1 (30.3–37.9) |  |
|  | 60–69 | 416 | 181.6 (5.6%) | 234.5 (22.7%) | 56.4 (49.6–63.1) |  |
| **Sex** |  |  |  |  |  | 0.631 |
|  | Men | 1,725 | 1,303.9 (39.9%) | 421.5 (40.8%) | 24.4 (22.1–26.7) |  |
|  | Women | 2,576 | 1,965.3 (60.1%) | 610.4 (59.2%) | 23.7 (21.8–25.6) |  |
| **Education** |  |  |  |  |  | 0.023 |
|  | No primary | 1,453 | 1,096.7 (33.5%) | 356.2 (34.6%) | 24.5 (22.1–26.9) |  |
|  | Primary | 1,038 | 783.1 (24.0%) | 254.7 (24.7%) | 24.5 (21.3–27.8) |  |
|  | Secondary | 1,430 | 1,124.4 (34.4%) | 305.2 (29.6%) | 21.4 (18.9–23.8) |  |
|  | Tertiary | 379 | 264.9 (8.1%) | 113.6 (11.0%) | 30.0 (24.2–35.8) |  |
| **Marital status** |  |  |  |  |  | <0.001 |
|  | Married/cohabiting | 2,598 | 1,980.1 (60.6%) | 618.1 (60.1%) | 23.8 (22.0–25.6) |  |
|  | Single | 916 | 760.9 (23.3%) | 155.2 (15.1%) | 16.9 (14.4–19.5) |  |
|  | Formerly married | 779 | 524.9 (16.1%) | 254.5 (24.8%) | 32.7 (28.2–37.1) |  |
| **Occupation** |  |  |  |  |  | <0.001 |
|  | Employed | 2,210 | 1,676.1 (51.3%) | 533.8 (51.8%) | 24.2 (22.1–26.2) |  |
|  | Unemployed | 1,514 | 1,105.0 (33.8%) | 409.1 (39.7%) | 27.0 (24.3–29.7) |  |
|  | Student/homemaker | 572 | 485.2 (14.9%) | 86.9 (8.4%) | 15.2 (12.1–18.3) |  |
| **Alcohol consumption** |  |  |  |  |  | 0.051 |
|  | No | 2,830 | 2,181.6 (66.7%) | 648.2 (62.8%) | 22.9 (21.2–24.6) |  |
|  | Yes | 1,471 | 1,087.5 (33.3%) | 383.7 (37.2%) | 26.1 (23.3–28.8) |  |
| **Daily smoking** |  |  |  |  |  | 0.713 |
|  | No | 3,952 | 3,000.6 (91.8%) | 950.9 (92.2%) | 24.1 (22.5–25.6) |  |
|  | Yes | 349 | 268.5 (8.2%) | 80.9 (7.8%) | 23.2 (18.6–27.7) |  |
| **Cooking oil type** |  |  |  |  |  | 0.791 |
|  | Vegetable/palm oil | 3,407 | 2,591.1 (81.6%) | 815.8 (81.0%) | 23.9 (22.3–25.6) |  |
|  | Animal/solid fats | 318 | 243.0 (7.7%) | 74.8 (7.4%) | 23.5 (18.4–28.6) |  |
|  | Other or none | 457 | 340.8 (10.7%) | 116.4 (11.6%) | 25.5 (21.2–29.7) |  |
| **Physical activity (vigorous)** |  |  |  |  |  | <0.001 |
|  | Active | 3,589 | 2,674.2 (81.8%) | 914.7 (88.6%) | 25.5 (23.8–27.1) |  |
|  | Inactive | 712 | 594.9 (18.2%) | 117.2 (11.4%) | 16.5 (13.5–19.4) |  |
| **Salt added to food** |  |  |  |  |  | 0.226 |
|  | Always | 1,273 | 948.1 (29.0%) | 324.6 (31.5%) | 25.5 (22.6–28.4) |  |
|  | Often | 363 | 271.7 (8.3%) | 91.1 (8.8%) | 25.1 (20.2–30.0) |  |
|  | Sometimes | 1,315 | 1,028.9 (31.5%) | 286.4 (27.8%) | 21.8 (19.3–24.3) |  |
|  | Rarely | 643 | 475.7 (14.6%) | 167.5 (16.2%) | 26.0 (22.3–29.7) |  |
|  | Never | 707 | 544.7 (16.7%) | 162.4 (15.7%) | 23.0 (19.3–26.6) |  |
| **High cholesterol** |  |  |  |  |  | <0.001 |
|  | No | 3,433 | 2,678.8 (92.9%) | 754.6 (82.4%) | 22.0 (20.5–23.5) |  |
|  | Yes | 367 | 205.8 (7.1%) | 160.8 (17.6%) | 43.9 (38.3–49.5) |  |
| **BMI category** |  |  |  |  |  | <0.001 |
|  | Normal weight | 2,678 | 2,210.1 (67.6%) | 467.8 (45.3%) | 17.5 (16.0–18.9) |  |
|  | Underweight | 268 | 214.0 (6.5%) | 53.5 (5.2%) | 20.0 (11.3–28.7) |  |
|  | Overweight | 835 | 570.7 (17.5%) | 264.3 (25.6%) | 31.7 (28.1–35.2) |  |
|  | Obese | 521 | 274.3 (8.4%) | 246.2 (23.9%) | 47.3 (41.3–53.3) |  |
| **Waist circumference (cm)** |  |  |  |  |  | <0.001 |
|  | Median (IQR) |  | 78.0 (73.0–85.0) | 85.0 (77.0–97.0) | — |  |
| **Diabetes (HbA1c)** |  |  |  |  |  | <0.001 |
|  | No diabetes (HbA1c <7%) | 3,407 | 2,639.0 (81.8%) | 767.8 (74.8%) | 22.5 (21.0–24.1) |  |
|  | Diabetes (HbA1c ≥7%) | 847 | 588.2 (18.2%) | 258.5 (25.2%) | 30.5 (26.3–34.8) |  |
| **Fruit and vegetable intake** | ≥5 servings/day | 1,048 | 790.2 (24.2%) | 257.8 (25.0%) | 24.6 (21.4–27.7) | — |
|  | <5 servings/day | 3,253 | 2,479.0 (75.8%) | 774.1 (75.0%) | 23.8 (22.2–25.3) |  |
| **Residence** |  |  |  |  |  | <0.001 |
|  | Rural | 2,534 | 1,986.7 (60.8%) | 547.6 (53.1%) | 21.6 (19.9–23.3) |  |
|  | Urban | 1,767 | 1,282.5 (39.2%) | 484.3 (46.9%) | 27.4 (24.8–30.1) |  |
| **Province** |  |  |  |  |  | <0.001 |
|  | Central | 340 | 258.3 (7.9%) | 81.5 (7.9%) | 24.0 (19.0–28.9) |  |
|  | Copperbelt | 725 | 558.1 (17.1%) | 166.4 (16.1%) | 23.0 (18.6–27.3) |  |
|  | Eastern | 518 | 408.1 (12.5%) | 110.3 (10.7%) | 21.3 (17.5–25.0) |  |
|  | Luapula | 400 | 313.3 (9.6%) | 86.3 (8.4%) | 21.6 (17.3–25.8) |  |
|  | Lusaka | 652 | 455.1 (13.9%) | 196.6 (19.1%) | 30.2 (26.2–34.1) |  |
|  | Muchinga | 321 | 230.0 (7.0%) | 90.9 (8.8%) | 28.3 (22.5–34.1) |  |
|  | Northern | 217 | 174.9 (5.4%) | 42.2 (4.1%) | 19.4 (14.0–24.9) |  |
|  | North-Western | 268 | 220.8 (6.8%) | 47.3 (4.6%) | 17.7 (12.8–22.5) |  |
|  | Southern | 595 | 417.0 (12.8%) | 177.5 (17.2%) | 29.9 (25.8–33.9) |  |
|  | Western | 266 | 233.5 (7.1%) | 32.9 (3.2%) | 12.4 (8.0–16.7) |  |

*a Weighted N values are shown; proportions are within-column percentages.*
*b Prevalence of hypertension (%) within each category with 95% confidence intervals.*
*p-values from chi-squared test for categorical variables and Wilcoxon rank-sum test for waist circumference.*
*Abbreviations: BMI, body mass index; HbA1c, glycated haemoglobin; IQR, interquartile range.*

### Multilevel Logistic Regression

To account for within-cluster correlation arising from the clustered survey design, generalized linear mixed models (GLMMs) with a PSU-level random intercept were fitted. The base model was:

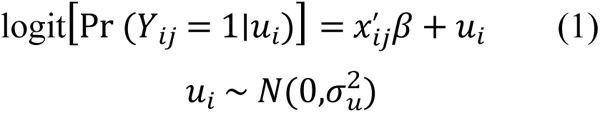

where *x*_i*j*_ represents the covariate vector, *β* the vector of fixed log-odds coefficients, and *u_i_* the PSU-specific random intercept. Adjusted odds ratios (AORs) were obtained by exponentiating *β*. Estimation used `lme4::glmer` (binomial family, Laplace approximation) with BOBYQA optimization and rescaled survey weights. Waist circumference was grand-mean centered; its AOR reflects a 1 cm increment above the sample mean.

### Model sequence

Four nested models were estimated. Model 0 (null) included only the PSU random intercept. Model I added all individual-level covariates. Model II included community-level variables (residence and province) only. The Full Model combined individual and community covariates. Model fit was compared using log-likelihood, Akaike Information Criterion (AIC), and Bayesian Information Criterion (BIC), with a likelihood ratio test comparing the null and Full GLMMs.

### Clustering diagnostics

The intraclass correlation coefficient (ICC) quantified the proportion of total variance attributable to between-PSU differences:

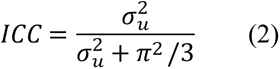

The median odds ratio (MOR) expressed between-cluster variability on the odds ratio scale:

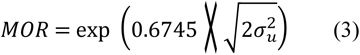

### Standard Logistic Regression (Sensitivity Analysis)

A conventional single-level logistic regression model (`stats::glm`, binomial family) with identical fixed effects to the Full Model was estimated as a sensitivity analysis to assess the influence of ignoring PSU clustering. Model adequacy was evaluated using McFadden pseudo-*R*^2^:

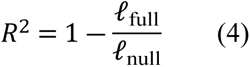

A likelihood ratio test against an intercept-only model was also performed. GLMM and GLM log-likelihoods are not directly comparable; therefore, no formal statistical comparison between the two modelling frameworks was conducted.

### Spatial Analysis

A georeferenced extract containing GPS coordinates, survey weights, PSU identifiers, province, and binary hypertension status was linked to GADM administrative boundaries (GeoJSON, WGS 84 / EPSG:4326). Survey-weighted hypertension prevalence for province *_p_*was computed as:

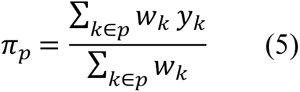

where *_wk_*and *_yk_*denote the survey weight and hypertension indicator for respondent *_k_*, respectively. Choropleth maps displayed both absolute prevalence and deviation from the national weighted mean.

Global spatial autocorrelation was assessed using Moran’s *_I_*, computed on individual-level binary outcomes with a row-standardized k-nearest-neighbour weights matrix (*_k_* = 8, great-circle distance; minor coordinate jitter applied with a fixed seed to resolve duplicates):

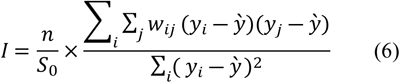

where:

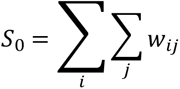

Statistical significance was evaluated under a normal approximation using `spdep::moran.test`. Local spatial clustering was characterized using Local Indicators of Spatial Association (LISA), classifying each observation as high-high, low-low, high-low, low-high, or not significant (*p* < 0.05), and overlaid on district boundaries. Spatial tests were exploratory and did not incorporate survey weights.

### Statistical Software

All analyses were conducted in R using the following packages: *haven*, *dplyr*, *tidyr*, *survey*, *lme4*, *sf*, *spdep*, *ggspatial*, *ggplot2*, *ggeffects*, and *patchwork*. Analysis scripts are available from the corresponding author upon reasonable request.

## Results

A total of 4,301 adults participated in the 2017 WHO STEPS survey in Zambia (Table 1), with 1,031.9 (24.0%) classified as hypertensive, giving an overall weighted prevalence of 24.0%. Most participants were women, 60.1% (n = 2,576), while men accounted for 39.9% (n = 1,725). The sample was largely young, with 35.3% (n = 1,520) aged 18 to 29 years and 34.8% (n = 1,498) aged 30 to 44 years. Age showed a strong association with hypertension (p < 0.001), increasing from 13.2% (95% CI: 11.4 to 15.0) in those aged 18 to 29 years to 56.4% (95% CI: 49.6 to 63.1) among those aged 60 to 69 years. Sex was not significantly associated with hypertension (p = 0.631), with similar prevalence in men at 24.4% and women at 23.7%. Education was significantly associated with hypertension (p = 0.023), with the highest prevalence among those with tertiary education at 30.0% (95% CI: 24.2 to 35.8) and the lowest among those with secondary education at 21.4% (95% CI: 18.9 to 23.8). Marital status was also significant (p < 0.001), with formerly married adults showing the highest prevalence at 32.7% (95% CI: 28.2 to 37.1) and single adults the lowest at 16.9% (95% CI: 14.4 to 19.5).

Among clinical and metabolic factors, high cholesterol was strongly associated with hypertension (p < 0.001), with prevalence of 43.9% (95% CI: 38.3 to 49.5) among those with high cholesterol compared to 22.0% (95% CI: 20.5 to 23.5) among those without. Obesity also showed a marked association (p < 0.001), with hypertension prevalence at 47.3% (95% CI: 41.3 to 53.3) versus 17.5% (95% CI: 16.0 to 18.9) in those with normal weight. Diabetes was significantly associated with hypertension (p < 0.001), with prevalence of 30.5% among individuals with HbA1c of 7% or higher. Urban residents had higher prevalence at 27.4% compared to rural residents at 21.6% (p < 0.001). At provincial level (p < 0.001), Lusaka recorded the highest prevalence at 30.2% while Western Province had the lowest at 12.4%. Physical activity showed an unexpected pattern, with the inactive group having lower prevalence at 16.5% compared to 25.5% among the active group (p < 0.001), likely reflecting reverse causation. Alcohol consumption, daily smoking, salt intake, cooking oil type, and fruit and vegetable intake were not statistically significant at the 5% level.

### Spatial distribution and global spatial autocorrelation of hypertension in Zambia

Figure 1 above illustrates the geographic distribution of hypertension prevalence across the ten provinces of Zambia, showing a clear spatial gradient with higher burden concentrated in the southern and central regions compared to the north and west. Lusaka Province recorded the highest prevalence at 29.9%, closely followed by Southern Province at 29.9%, while Muchinga Province also showed elevated levels at 28.3%. In contrast, Western Province had the lowest prevalence nationally at 12.4%, followed by North Western Province at 17.7% and Northern Province at 19.5%.

**Fig. 1.**
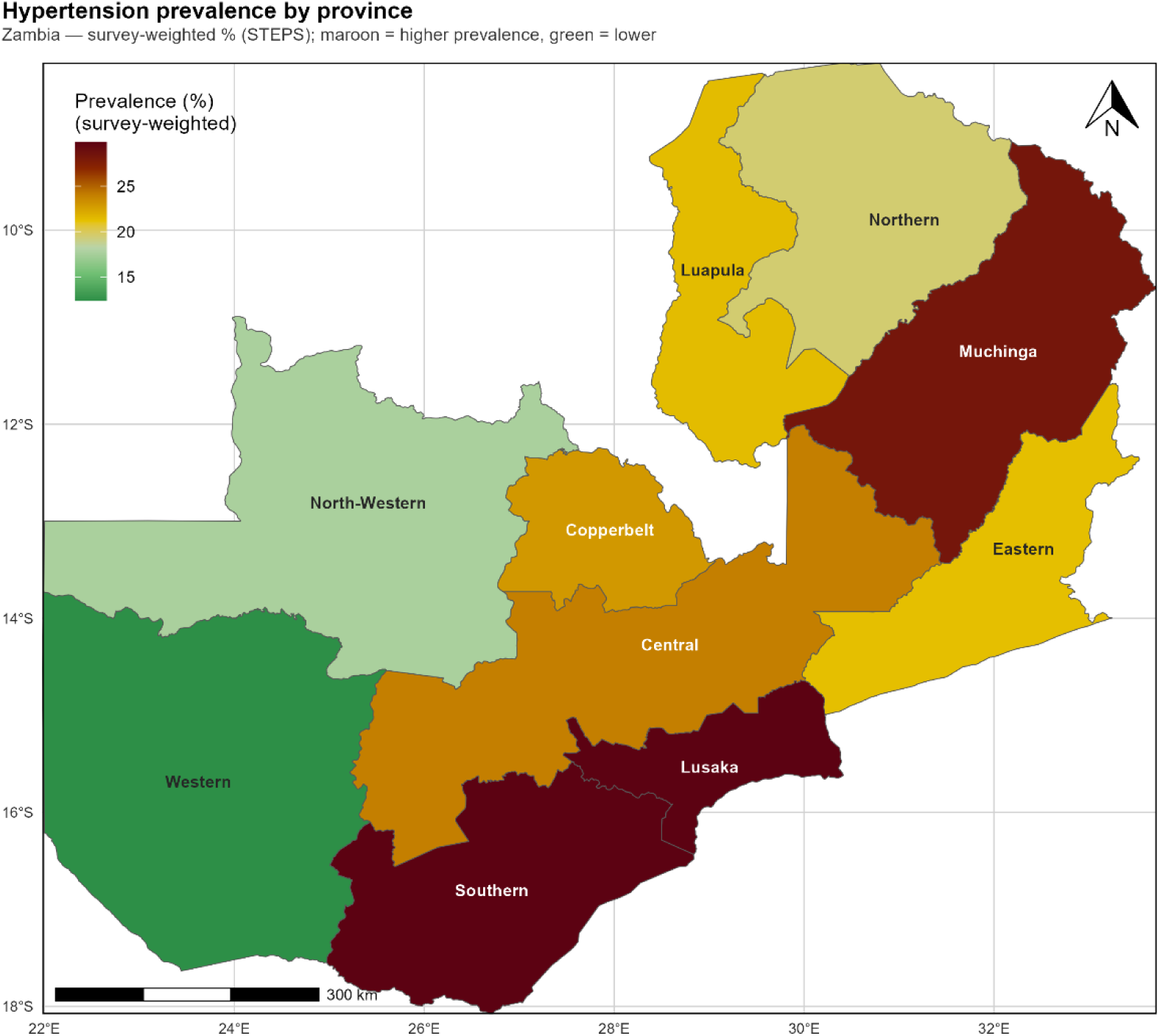
Map of Zambia showing the prevalence of High blood pressure by provincial level. The base layer of the map was obtained from the Database of Global Administrative Areas (GADM) version 4.1 (https://gadmorg/download_country.html). GADM data is freely available for academic and non-commercial use and is compatible with the CC BY 4.0 license. prevalence of High Blood Pressure by provincial level data were obtained from the 2017 STEPS survey.

Central, Eastern, Copperbelt, and Luapula provinces formed a mid-range cluster with prevalence estimates between 21% and 24%. Overall, this provincial pattern suggests a meaningful geographic variation in hypertension risk in Zambia, with higher prevalence in more urbanised and economically developed areas, consistent with broader urbanisation and lifestyle transition patterns observed in other sub-Saharan African settings.

Global Moran’s I showed a small but statistically significant level of spatial clustering in hypertension among 4003 participants (I = 0.0247, z = 3.42, p < 0.001). Individuals with hypertension were slightly more likely to be located near others with hypertension, and the same pattern was observed for normotensive individuals, compared with what would be expected under complete spatial randomness. However, the magnitude of clustering was weak, as the observed Moran’s I was only marginally higher than the null expectation (−0.00025), indicating limited practical importance at the individual level across Zambia.

The observed spatial pattern reflects both the underlying distribution of sampled locations and the nature of hypertension risk across the population. The spatial weights matrix included 271 disconnected components, primarily arising from naturally sparse or geographically isolated GPS points, which is common in nationally representative survey data and contributes to weaker global spatial dependence because such locations are not strongly linked within broader spatial networks. Minor coordinate jitter used to manage duplicate locations introduced only negligible positional variation and does not affect the substantive interpretation of spatial structure. Overall, hypertension exhibits statistically detectable but spatially diffuse clustering, consistent with a modest and locally varying geographic influence rather than strong countrywide spatial dependence. These localized patterns are further detailed through LISA cluster maps presented in Figure 2

**Fig. 2.**
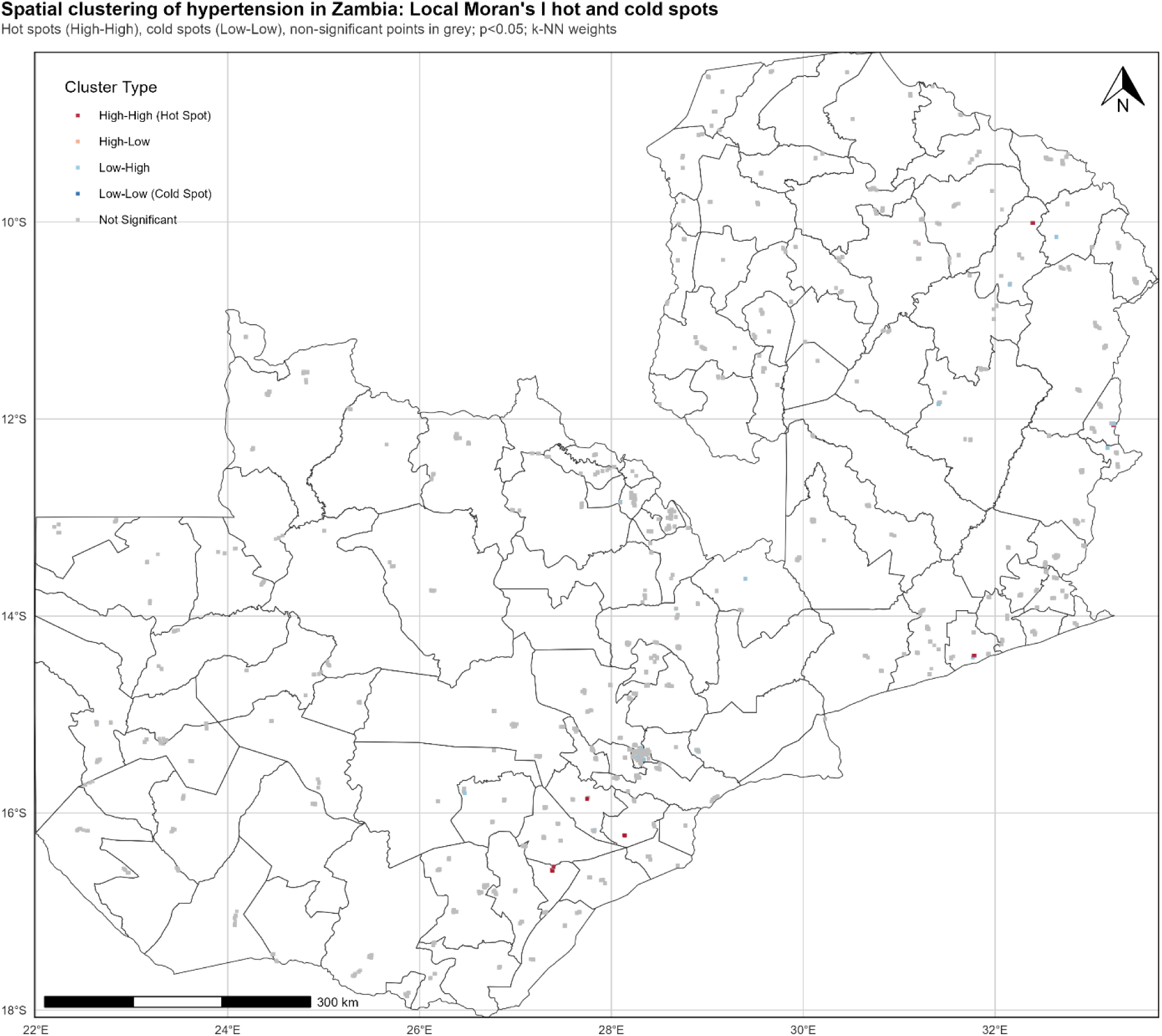
Map of Zambia showing the spatial clustering of High blood pressure in Zambia: Local Morans I of Hotspots and cold spots. The base layer of the map was obtained from the Database of Global Administrative Areas (GADM) version 4.1 (https://gadmorg/download_country.html). GADM data is freely available for academic and non-commercial use and is compatible with the CC BY 4.0 license. Spatial clustering of High Blood pressure in Zambia: Local Morans I of Hotspots and cold spots obtained from the 2017 WHO STEPS Survey

### Multilevel Logistic Regression Models for factors associated Hypertension

Table 2 below presents results from four nested multilevel logistic regression models assessing hypertension. The null model showed a between-cluster variance of 0.2049 and an intraclass correlation coefficient (ICC) of 5.9%, indicating that about 6% of hypertension variation was attributable to differences between primary sampling units before adjustment. The median odds ratio (MOR) was 1.54, meaning an individual’s odds of hypertension could be 1.54 times higher simply due to cluster location. This confirmed meaningful clustering and justified a multilevel approach.

**Table 2.** Multilevel logistic regression models for hypertension among adults in Zambia (N = 3,474)

| Characteristic | Level | Null Model | Model I: Individual-level | Model II: Community-level | Full Model (All) |
| --- | --- | --- | --- | --- | --- |
|  |  | AOR (95% CI) | AOR (95% CI) | AOR (95% CI) | AOR (95% CI) |
| Age group (years) |  |  |  |  |  |
|  | 18–29 (Ref.) | — | 1.00 | — | 1.00 |
|  | 30–44 | — | 1.46 (1.14 – 1.87)** | — | 1.49 (1.16 – 1.91)** |
|  | 45–59 | — | 2.75 (2.09 – 3.62)*** | — | 2.86 (2.17 – 3.77)*** |
|  | 60–69 | — | 6.75 (4.84 – 9.41)*** | — | 6.92 (4.95 – 9.66)*** |
| Sex |  |  |  |  |  |
|  | Men (Ref.) | — | 1.00 | — | 1.00 |
|  | Women | — | 0.64 (0.52 – 0.79)*** | — | 0.64 (0.52 – 0.79)*** |
| Education |  |  |  |  |  |
|  | Tertiary (Ref.) | — | 1.00 | — | 1.00 |
|  | No primary | — | 0.84 (0.60 – 1.17) | — | 0.93 (0.66 – 1.30) |
|  | Primary | — | 0.85 (0.61 – 1.19) | — | 0.89 (0.63 – 1.24) |
|  | Secondary | — | 0.86 (0.63 – 1.17) | — | 0.87 (0.63 – 1.19) |
| Marital status |  |  |  |  |  |
|  | Married/cohabiting (Ref.) | — | 1.00 | — | 1.00 |
|  | Single | — | 1.33 (1.02 – 1.73)* | — | 1.34 (1.03 – 1.75)* |
|  | Formerly married | — | 1.15 (0.91 – 1.44) | — | 1.14 (0.90 – 1.43) |
| Occupation |  |  |  |  |  |
|  | Employed (Ref.) | — | 1.00 | — | 1.00 |
|  | Unemployed | — | 1.08 (0.89 – 1.31) | — | 1.04 (0.84 – 1.27) |
|  | Student/homemaker | — | 0.76 (0.55 – 1.03) | — | 0.75 (0.55 – 1.03) |
| Alcohol consumption |  |  |  |  |  |
|  | No (Ref.) | — | 1.00 | — | 1.00 |
|  | Yes | — | 1.12 (0.93 – 1.35) | — | 1.14 (0.94 – 1.38) |
| Daily smoking |  |  |  |  |  |
|  | No (Ref.) | — | 1.00 | — | 1.00 |
|  | Yes | — | 0.79 (0.57 – 1.11) | — | 0.80 (0.58 – 1.12) |
| Cooking oil type |  |  |  |  |  |
|  | Vegetable/palm oil (Ref.) | — | 1.00 | — | 1.00 |
|  | Animal/solid fats | — | 1.11 (0.81 – 1.53) | — | 1.18 (0.81 – 1.72) |
|  | Other or none | — | 1.00 (0.75 – 1.32) | — | 1.05 (0.78 – 1.41) |
| **Physical activity (vigorous)** |  |  |  |  |  |
|  | Active (Ref.) | — | 1.00 | — | 1.00 |
|  | Inactive | — | 0.89 (0.69 – 1.16) | — | 0.93 (0.71 – 1.21) |
| **Salt added to food** |  |  |  |  |  |
|  | Never (Ref.) | — | 1.00 | — | 1.00 |
|  | Always | — | 1.10 (0.83 – 1.45) | — | 1.02 (0.77 – 1.36) |
|  | Often | — | 1.23 (0.85 – 1.78) | — | 1.21 (0.83 – 1.76) |
|  | Sometimes | — | 1.10 (0.84 – 1.44) | — | 1.12 (0.85 – 1.48) |
|  | Rarely | — | 1.22 (0.90 – 1.65) | — | 1.24 (0.91 – 1.69) |
| **High cholesterol** |  |  |  |  |  |
|  | No (Ref.) | — | 1.00 | — | 1.00 |
|  | Yes | — | 1.44 (1.10 – 1.87)** | — | 1.40 (1.07 – 1.82)* |
| **BMI category** |  |  |  |  |  |
|  | Normal weight (Ref.) | — | 1.00 | — | 1.00 |
|  | Underweight | — | 1.12 (0.74 – 1.71) | — | 1.12 (0.74 – 1.71) |
|  | Overweight | — | 1.71 (1.36 – 2.15)*** | — | 1.65 (1.30 – 2.08)*** |
|  | Obese | — | 2.46 (1.77 – 3.43)*** | — | 2.34 (1.67 – 3.26)*** |
| **Diabetes (HbA1c)** |  |  |  |  |  |
|  | No diabetes, HbA1c <7% (Ref.) | — | 1.00 | — | 1.00 |
|  | Diabetes, HbA1c ≥7% | — | 1.37 (1.07 – 1.77)* | — | 1.35 (1.04 – 1.75)* |
| **Fruit and vegetable intake** |  |  |  |  |  |
|  | ≥5 servings/day (Ref.) | — | 1.00 | — | 1.00 |
|  | <5 servings/day | — | 0.84 (0.66 – 1.06) | — | 0.86 (0.68 – 1.09) |
| **Waist circumference (cm)** |  |  |  |  |  |
|  | Per 1 cm increase (grand-mean centred) | — | 1.02 (1.01 – 1.03)*** | — | 1.02 (1.01 – 1.03)*** |

| Residence |  |  |  |  |  |
| --- | --- | --- | --- | --- | --- |
|  | Rural (Ref.) | — | — | 1.00 | 1.00 |
|  | Urban | — | — | 1.42 (1.16 – 1.74)*** | 1.20 (0.95 – 1.50) |
| Province |  |  |  |  |  |
|  | Central (Ref.) | — | — | 1.00 | 1.00 |
|  | Copperbelt | — | — | 0.78 (0.53 – 1.15) | 0.89 (0.59 – 1.33) |
|  | Eastern | — | — | 0.86 (0.59 – 1.27) | 0.81 (0.54 – 1.22) |
|  | Luapula | — | — | 0.89 (0.58 – 1.36) | 0.93 (0.58 – 1.49) |
|  | Lusaka | — | — | 1.13 (0.77 – 1.64) | 1.07 (0.71 – 1.59) |
|  | Muchinga | — | — | 1.24 (0.82 – 1.87) | 1.23 (0.78 – 1.93) |
|  | Northern | — | — | 0.78 (0.49 – 1.24) | 0.78 (0.45 – 1.33) |
|  | North-Western | — | — | 0.65 (0.42 – 1.03) | 0.73 (0.46 – 1.18) |
|  | Southern | — | — | 1.28 (0.89 – 1.84) | 1.16 (0.79 – 1.71) |
|  | Western | — | — | 0.46 (0.28 – 0.75)** | 0.48 (0.29 – 0.81)** |
| Model fit statistics |  |  |  |  |  |
|  | N | 3,474 | 3,474 | 3,474 | 3,474 |
|  | Log-likelihood | −2016.17 | −1746.61 | −1989.69 | −1733.61 |
|  | AIC | 4036.34 | 3551.23 | 4003.38 | 3545.23 |
|  | BIC | 4048.65 | 3729.67 | 4077.22 | 3785.20 |
|  | Var(PSU) | 0.2049 | 0.0861 | 0.1105 | 0.0594 |
|  | ICC | 0.0586 | 0.0255 | 0.0325 | 0.0177 |
|  | MOR | 1.54 | 1.32 | 1.37 | 1.26 |

AOR, adjusted odds ratio; CI, confidence interval; ICC, intraclass correlation coefficient; MOR, median odds ratio; PSU, primary sampling unit; Var(PSU), between-cluster variance.
Reference categories denoted as 1.00 (Ref.); — indicates variable not included in that model.
Null Model: intercept-only (no predictors). Model I: individual-level predictors only. Model II: community-level predictors only. Full Model: all individual- and community-level predictors.
Significance: \*\*\* $p < 0.001$ ; \*\* $p < 0.01$ ; \* $p < 0.05$ .
$ICC = Var(PSU) / [Var(PSU) + \pi^2/3]$ ; $MOR = \exp(0.6745 \times \sqrt{[2 \times Var(PSU)]})$ .

In Model I (individual-level factors only), clustering reduced substantially, with variance dropping to 0.0861, ICC to 2.6%, and MOR to 1.32, showing that individual characteristics explained much of the between-cluster variation. Model II (community-level factors only) showed ICC = 3.3% and MOR = 1.37, indicating a moderate contribution of contextual factors. The Full Model provided the best fit (AIC = 3,545.23; BIC = 3,785.20; log-likelihood = 1,733.61), with residual variance reduced to 0.0594, ICC = 1.8%, and MOR = 1.26, suggesting most clustering was explained when both individual and community factors were included.

In the Full Model, age remained the strongest predictor: compared to ages 18–29 years, those aged 30–44 had higher odds (AOR 1.49, 95% CI: 1.16–1.91), 45–59 years (AOR 2.86, 95% CI: 2.17–3.77), and 60–69 years (AOR 6.92, 95% CI: 4.95–9.66), all p < 0.001. Women had lower odds than men (AOR 0.64, 95% CI: 0.52–0.79, p < 0.001). Single individuals had higher odds than married/cohabiting adults (AOR 1.34, 95% CI: 1.03–1.75, p < 0.05). Obesity (AOR 2.34, 95% CI: 1.67–3.26) and overweight (AOR 1.65, 95% CI: 1.30–2.08) were significant predictors (both p < 0.001), as was waist circumference (AOR 1.02 per cm, 95% CI: 1.01–1.03, p < 0.001). High cholesterol (AOR 1.40, 95% CI: 1.07–1.82) and diabetes (AOR 1.35, 95% CI: 1.04–1.75) were also significant (p < 0.05). Urban residence was significant in Model II (AOR 1.42, 95% CI: 1.16–1.74, p < 0.001) but not in the Full Model (AOR 1.20, 95% CI: 0.95–1.50), suggesting attenuation after individual adjustment. Western Province consistently showed lower odds than Central Province in Model II (AOR 0.46, 95% CI: 0.28–0.75) and the Full Model (AOR 0.48, 95% CI: 0.29–0.81), both p < 0.01. Other factors (education, occupation, alcohol, smoking, salt intake, physical activity, cooking oil, fruit/vegetable intake) were not significant.

## Discussion

This study examined the spatial distribution and multilevel determinants of hypertension among Zambian adults using the 2017 WHO STEPS survey. One in four adults (24.0%) was hypertensive, with the burden concentrated among older adults, men, single individuals, and those living with obesity, elevated waist circumference, raised cholesterol, and diabetes. Spatially, hypertension was most concentrated in Lusaka and Southern Province, while Western Province consistently showed lower risk. Although spatial clustering was statistically significant, its magnitude was modest, and the multilevel analysis showed that most between community variation was explained once individual level characteristics were accounted for.

The 24.0% prevalence aligns closely with the pooled estimate of 27.09% reported in a recent systematic review and meta analysis of 52 community based studies in sub Saharan Africa^19^. It also mirrors the 24.5% found in the 2015 Kenya STEPS survey of 4,485 adults^20^. However, the figure is lower than the 34.8% reported from urban Lusaka over a decade ago^10^ and the 32.3% observed in urban Kitwe ^21^, likely reflecting the nationally representative STEPS design, which captures a younger and more rural population than earlier clinic or city level studies. In this context, the observed prevalence still represents a substantial public health burden, particularly given that globally, hypertension has shifted toward low and middle income countries, where only one in three affected individuals is aware of their status and fewer than 8% have blood pressure controlled^22^.

Age was the strongest predictor of hypertension, with risk increasing clearly with each decade, a pattern consistently reported in African STEPS studies. In Rwanda, adults aged 60 to 69 were 5.7 times more likely to be hypertensive than the youngest group^23^, and in Malawi, prevalence rose from 21.4% in 25 to 34 year olds to 59.2% in 55 to 64 year olds^24^. However, our finding that women had lower odds after full adjustment differs from some regional evidence; for example, in Senegal, women had higher prevalence than men (47.9% versus 41.7%)^25^, while a systematic analysis reported that men had 1.80 times higher odds of hypertension across Africa^19^. The lower female risk in our younger sample likely reflects the protective effect of premenopausal oestrogen on vascular tone and sodium handling, although this advantage probably reduces with age (Xiang et al., 2021).

Being single was linked to higher odds of hypertension compared to being married or cohabiting, consistent with broader cardiovascular evidence that partnerships offer psychosocial and material support, including better care seeking and lower chronic stress (Fan et al., 2025; Laurito et al., 2025; Conklin & Guo et al., 2024). Both overweight and obesity were strongly associated with hypertension, as was increasing waist circumference. A meta analysis reported that abdominal obesity was linked to 2.86 times higher odds of hypertension in sub Saharan Africa^19^, while a large harmonised dataset across 13 African countries found obese individuals were 2.5 times more likely to be hypertensive, with odds rising from 2.0 in younger adults to 8.8 in older groups^26^. The role of central fat is well understood, as excess visceral fat activates the renin angiotensin aldosterone system, promotes insulin resistance, and drives chronic low grade inflammation^27^.

Diabetes and elevated cholesterol were each independently associated with hypertension. This co-occurrence of cardiometabolic risk factors is well documented across Africa, with evidence showing that more than one in four Zambian adults had three or more NCD risk factors at the same time^28^. The lack of independent associations for behavioural factors such as smoking, alcohol use, salt intake, and physical activity in the adjusted model differs from broader evidence, where these factors are often significant predictors^19^. The lower prevalence observed among physically inactive individuals is likely due to reverse causation, where those already unwell reduce vigorous activity, a limitation that cross sectional designs cannot fully address.

Spatially, hypertension was more common in the south and central parts of the country, areas with higher urbanisation, more sedentary work, and greater reliance on processed foods. Western Province consistently showed lower risk, echoing earlier findings that physically demanding rural livelihoods and less commercialised food environments may be protective^11^. Similar geographic patterns have been reported elsewhere, with higher prevalence in more urbanised areas and lower levels in rural settings^18^, and clustering observed near major transport routes and areas with more amenities (Burns et al., 2021). Our multilevel results also showed community level clustering in the null model, which largely reduced after adjusting for individual factors, suggesting that these geographic differences are mainly due to population characteristics rather than place based effects.

These findings carry direct implications for Sustainable Development Goal Target 3.4, which mandates a one third reduction in premature NCD mortality by 2030. Progress has been slowest in low and middle income countries ^15^, and blood pressure control remains below 10% in rural settings across the region ^22^. For Zambia, the combination of high prevalence, very low awareness, and absence of routine screening represents a missed opportunity for early intervention.

### Policy Implications

From a policy perspective, these findings support integrating blood pressure screening into existing primary care platforms, particularly in Lusaka and Southern Province where the burden is highest. The WHO HEARTS technical package and community health worker led models have shown promise in similar settings across Bangladesh, Pakistan, and Kenya^29^. Population level salt reduction policies and alcohol taxation, identified as among the most cost effective NCD interventions globally (Watkins et al., 2022), are directly relevant to the Zambian context. For research, longitudinal follow up of the STEPS cohort would clarify temporal relationships between behavioural risk factors and hypertension onset. Province level studies with finer spatial resolution would better characterise the environmental and contextual determinants underlying the clustering observed here.

### Strengths and Limitations

Strengths of this study include the use of a nationally representative WHO STEPS dataset with standardised blood pressure measurement, the application of multilevel modelling that accounts for the hierarchical data structure, and the incorporation of spatial analysis to identify geographic clustering.

Limitations include the cross sectional design, which prevents causal inference and cannot resolve the directionality of associations such as that between physical activity and hypertension. Self reported behavioural variables such as salt intake and alcohol consumption are subject to social desirability bias. The spatial analysis was constrained to provincial level aggregation, which may mask finer grained within province variation. Additionally, the STEPS survey did not collect data on medication use or awareness, limiting our ability to characterise the treatment cascade.

### Conclusion

Hypertension affects approximately one in four Zambian adults and is driven primarily by advancing age, male sex, single marital status, overweight and obesity, central adiposity, diabetes, and raised cholesterol. Spatial clustering in southern and central provinces reflects differential urbanisation and nutritional transition. Strengthening blood pressure screening, integrating cardiometabolic risk management within primary care, and implementing population level dietary and alcohol policies represent the most promising pathways toward meeting SDG Target 3.4 in Zambia.

## Data Availability

The data used in this study are publicly available from the World Health Organization (WHO) STEPwise Approach to NCD Risk Factor Surveillance (STEPS) program. The 2017 Zambia STEPS dataset can be requested from the WHO NCD Microdata Repository (https://extranet.who.int/ncdsmicrodata/index.php/catalog). Access to the dataset requires registration and approval from WHO, in line with their data sharing policies. The authors did not have any special access privileges to the data beyond those granted to other researchers. All derived datasets and statistical code used for analysis can be obtained from the corresponding author upon reasonable request.

## Ethics Statement

The 2017 Zambia World Health Organization STEPwise Approach to Surveillance (WHO STEPS) survey was conducted in accordance with internationally accepted ethical standards for research involving human participants. Ethical approval for the original survey was obtained from the relevant national ethics bodies in Zambia and the World Health Organization Ethics Review Committee. Written informed consent was obtained from all participants prior to data collection.

This study involved secondary analysis of anonymised WHO STEPS survey data obtained from the WHO NCD Microdata Repository. The dataset is publicly available upon request and contains no personally identifiable information. No direct contact with human participants occurred during this study. Therefore, additional ethical approval was not required for the present analysis.

## Data Availability

The data underlying the results presented in this study are available from the World Health Organization NCD Microdata Repository upon reasonable request and approval. The Zambia 2017 WHO STEPS survey dataset can be accessed through the WHO NCD Microdata Repository website.

